# Trauma-spectrum symptoms among the Italian general population in the time of the COVID-19 outbreak

**DOI:** 10.1101/2020.06.01.20118935

**Authors:** Rodolfo Rossi, Valentina Socci, Dalila Talevi, Cinzia Niolu, Francesca Pacitti, Antinisca Di Marco, Alessandro Rossi, Alberto Siracusano, Giorgio Di Lorenzo, Miranda Olff

## Abstract

**Background:** Recent evidence showed substantial negative mental health outcomes associated with the current COVID-19 pandemic, including trauma-related symptoms although the effects on the Italian population who were subjected to unprecedented nationwide lockdown measure remains unknown. The Global Psychotrauma Screen (GPS) is a brief instrument designed to assess a broad range of trauma-related symptoms with no available validation in the Italian population.

**Aims:** This study aimed at examining the factor structure of the Italian version of the GPS in a general population sample exposed to the COVID-19 pandemic and at evaluating trauma-related symptoms in the Italian population in the context of specific COVID-19 related risk factors associated with the implementation of lockdown measures and social distancing.

**Methods:** Cross-sectional web-based observational study, as part of a long-term monitoring programme of mental health outcomes in the general population. 18147 participants completed a self-report online questionnaire to collect key demographic data and to evaluate trauma-related symptoms using the GPS, PHQ-9, GAD-7, ISI and PSS. Validation analyses included both exploratory and confirmatory factor analysis, and correlation analyses.

**Results:** Exploratory factor analyses supported both a two-factor and a three-factor model. Confirmatory factor analysis showed that a one-factor solution that was used as a baseline comparison showed acceptable fit indices, the two-factor solution showed good fit indices, but the best fitting model was a three-factor solution, with Negative Affect (symptoms of depressed mood, anxiety, irritability), core Post-traumatic Stress Symptoms (PTSS) (avoidance, re-experiencing, hyperarousal and insomnia) and Dissociative symptoms. GPS Risk factors as well as specific COVID-19 related stressful events, were associated with GPS total as well as the three factor scores.

**Conclusions:** Our data suggest that a wide range of trauma-spectrum symptoms were reported by a large Italian sample during the COVID-19 pandemic. The GPS symptoms clustered best in three factors: Negative Affect symptoms, Core PTSS, and Dissociative symptoms. In particular high rates of core PTSS and negative affect symptoms were associated with the COVID-19 pandemic in Italy and should be routinely assessed in clinical practice.

## 1. Introduction

The 2019 coronavirus (COVID-19) pandemic is a global health emergency with serious impact on public health, including mental health (Brooks et al., 2020). At the time this paper was written, the pandemic was affecting Europe and Italy in the first place, leading to the implementation of unprecedented preventive measures aimed at containing the spread of infections. By the first decade of March 2020, lockdown measures were imposed on the Italian territory by the Government. People were asked to stay at home and socially isolate themselves to prevent being infected.

It has been previously shown that health emergencies such as epidemics can lead to detrimental and long-lasting psychosocial consequences. At the individual level, epidemics are associated with a wide range of psychiatric comorbidities including anxiety, panic, depression and trauma-related disorders (Tucci et al., 2017). The psychosocial impact of health emergences seems to be even higher during quarantine measures (Brooks et al., 2020). Quarantine has been associated with high stress levels (DiGiovanni et al., 2004), depression (Hawryluck et al., 2004) irritability and insomnia (Lee et al., 2005). Further, being quarantined was a predictor of acute stress (Bai et al., 2004) and trauma-related (Wu et al., 2009) disorders, particularly in specific at-risk populations such as health-care workers. Trauma-related symptoms may persist long after the actual emergency has ended, thus representing a long-lasting threat to mental health. In the context of the current COVID-19 pandemic, an online survey on 1210 Chinese subjects during the initial stage of the disease reported moderate to severe stress levels in 8,1% of the respondents (Wang et al., 2020). A month after the COVID-19 outbreak, the prevalence of PTSD symptoms was found to be 7% in a sample of 285 participants from China hardest-hit areas (Liu et al., 2020). In a nationwide survey including more than 50.000 Chinese respondents, almost 35% of the participants reported trauma-related distress symptoms (Qiu et al., 2020).

Demographic risk factors for mental health outcomes such as female gender have been well established (Olff et al., 2017). Liu et al., (2020) also reported significant higher posttraumatic stress symptoms (PTSS) in women, particularly in the domains of re-experiencing, alterations in cognition and mood and hyperarousal (Liu et al., 2020). Also in the context of COVID-19 female and young adults showed significantly higher psychological distress (Qiu et al., 2020). Furthermore, the incidence of anxiety and stress disorders was high in front-line medical staff (Huang et al., 2020). Among Chinese health-care workers, women, nurses, front-line workers and those who worked in the hardest-hit areas were at higher risk of negative mental health outcomes including anxiety, insomnia and distress (Lai et al., 2020), while social support has been shown to positively affect stress, anxiety and self-efficacy levels in medical staff (Xiao et al., 2020). In addition to the lack of social support other well known risk factors for posttraumatic stress reactions include social economic status, a history of psychotrauma or psychiatric diagnoses, or specific event characteristics (Olff, 2017)

No study to date has investigated the range of trauma-related symptoms and associated risk factors of the COVID-19 pandemic in the Italian population. Considering the implementation of the strict lockdown and social distancing measures imposed on the entire national territory it is expected to have a significant impact on a wide range of mental health symptoms.

Recently, the Global Collaboration on Traumatic Stress (https://www.global-psychotrauma.net) developed the Global Psychotrauma Screen (GPS) (Schnyder et al., 2017; Olff et al., 2020), a brief instrument designed to assess trauma-related symptoms as well as associated risk and protective factors. GPS consists of 17 items encompassing different symptoms domains, such as PTSD and complex PTSD, depression, anxiety, sleep problems, self-injurious behaviour, dissociation, other physical, emotional or social problems, substance abuse; and 5 risk or protective factors such as other stressful events, childhood trauma, history of mental illness, social support and resilience. Preliminary data suggested good internal reliability as well as concurrent validity with instruments measuring trauma-related domains (Olff et al., 2020). The instrument therefore has the potential to represent a brief screening measure of a range of psychotrauma-related symptoms not limited to core PTSD symptoms.

This study aimed at examining the endorsement of symptoms and the factor structure of the Italian version of the GPS in a general population sample exposed to the COVID-19 pandemic and at evaluating mental health risk factors including specific COVID-19 related risk factors associated with the implementation of lockdown measures and social distancing.

## 2. Methods

### 2.1 Study design

This study is a part of a long-term monitoring programme of mental health outcomes in the general population (Rossi et al., 2020a, 2020b). Approval for this study was obtained from the local IRB at University of L’Aquila. On-line consent was obtained from the participants. Participants were allowed to terminate the survey at any time they desired. The survey was anonymous, and confidentiality of information was assured.

At three weeks after the beginning of the lockdown, a survey was conducted among a self-selected sample. Every person living in Italy ≥ 18 years was eligible. Participants were invited using sponsored social network advertisement together with a snowball recruiting technique.

Data on mental health were collected between March 27th and April 6^th^, 2020 using an on-line questionnaire developed using the free software Google Forms®. The investigated timeframe corresponds to the contagion peak in Italy, according to epidemiological data confirmed by the World Health Organization (World Health Organization, 2020).

### 2.2 Study Sample

A total of 18147 individuals completed the questionnaire, of which 14447 (79.6%) women, median age was 38 (IQR = 23). Because of the web-based design, no response rate could be estimated as it was not possible to estimate how many persons were reached by social network advertisement.

### 2.3 Measures

#### 2.3.1 Global Psychotrauma Screen

The Global Psychotrauma Screen (GPS) (Olff et al., 2020) is a 22 self-report instrument with yes/no answers that covers both stress-related symptoms and risk and protective factors. Symptoms investigated are (17 items): post-traumatic stress symptoms, depression, sleep problems, dissociation, dysfunctional coping strategies including substance abuse and self-harm, and other physical, emotional or social problems. Risk and protective factors are (5 items): other stressful events, childhood trauma, history of mental illness, social support and psychological resilience. The following scores were derived from the GPS: 1) “GPS symptoms” (GPS-Sym): sum of all 17 symptoms items 2) “GPS risk/protection index” (GPS-RP): sum of the 5 risk/protection items.

#### 2.3.2 Patient Health Questionnaire

Depressive symptoms were assessed using the 9-item Patient Health Questionnaire (PHQ-9). PHQ-9 comprises nine depressive symptoms, rated on a 4-point likert scale, range 0–27 (Spitzer et al., 1999). The total score has been taken into consideration as continuous variable. PHQ-9 is a widely used instrument in epidemiological research as depression screener. In our sample, internal consistency was α = 0.87.

#### 2.3.3 Generalized Anxiety Disorder questionnaire

Anxiety symptoms were assessed using the 7-item Generalized anxiety disorder questionnaire (GAD-7), rated on a 4-point likert scale, range 0–21 (Spitzer et al., 2006). The total score has been taken into consideration as continuous variable. GAD-7 is a widely used instrument in epidemiological research as anxiety screener. In our sample, internal consistency was α = 0.91.

#### 2.3.4 Insomnia severity index

The Insomnia Severity Index (ISI) is a 7-item self-report questionnaire assessing the nature, severity and impact of insomnia, on 5-point likert scale, range 0–28, with higher scores indicating higher severity of insomnia symptoms (Bastien et al., 2001; Castronovo et al., 2016). The total score has been taken into consideration as continuous variable. ISI is a widely used instrument to evaluate sleep disorders. In our sample, internal consistency was α = 0.90.

#### 2.3.5 Perceived stress scale

The 10-item Perceived Stress Scale (PSS) was used to assess subjectively perceived stress, on a 5-point likert scale, range 0–50 (Mondo et al., 2019). Internal consistency in our sample was α = 0.87.

#### 2.3.6 COVID-19 related risk factors

In this study, the impact of COVID-19 pandemic and related lockdown measures were addressed exploring 1) lockdown status, i.e. lockdown as imposed by the government, being under quarantine because infected or exposed to an infected person, hospitalized; 2) any change in working activity due to the COVID-19 pandemic, i.e. working as usual, working from home, working activity discontinued, working more than usual; 3) having a loved one infected, hospitalized or deceased due to COVID-19; 4) Any other problem event due to pandemic or lockdown, including financial, own’s or a loved one’s health, work/study, relational, housing or caregiving problems. Finally, gender, age, job, education, and region of residence were collected as demographic variables.

### 2.4 Analytic Plan

Firstly, an **exploratory factor analysis (EFA)** on the GPS symptoms was performed using principal axis factoring (PAF), followed by oblique promax rotation, in order to allow correlations between factors. Standard factor analyses are based on a matrix of Pearson’s correlations, and assume that the variables are continuous and follow a multivariate normal distribution. Because the GPS has 0/1 response items, a factor analysis based on tetrachoric correlations was performed.

Factor retention was based on Eigenvalue, screeplot inspection as well as Aikake’s and Bayesian Information Criteria (AIC and BIC) values comparison. The internal consistency of factors was examined using Cronbach’s alpha, with the threshold of .7 used to indicate acceptable reliability. Furthermore, item-test and item-rest correlations were estimated as measures of reliability. Barlett’s sphericity test and Kaiser–Meyer–Olkin (KMO) measure were inspected to ascertain data suitability for factor analysis.

In order to assess convergent validity with depression and anxiety symptoms, Pearson correlation between GPS-Sym, GPS-Sym resulting factor sub-scores with PHQ GAD and ISI were performed. Divergent validity was assessed by correlation with the PSS.

Secondly, a **confirmatory factor analysis (CFA)** was conducted on the models produced by EFA. Although it is generally desirable to perform EFA and CFA on two different samples, in this case CFA has been performed with the scope of assessing which model resulted from EFA had better goodness-of-fit indices (GOF).

Model parameter appraisal used maximum likelihood (ML) estimation. Multiple GOF indices including chi-square test, Comparative Fit Index (CFI), Standardized Root-Mean-Square Residual (SRMR), and Root-Mean-Square Error of Approximation (RMSEA) evaluated model fit. Using a range of indices ensures robust assessment of model fit.

In order to assure equal GOF appraisal, CFA models were not modified according to modification indices.

Subsequently, we explored the role of GPS-RP as an independent variable for a number of different outcomes, including GPS-Sym, and the resulting GPS factors.

After establishing the best fitting factor structure of the GPS-Sym, GPS-RP was introduced as independent variable in a panel of seemingly unrelated regression models on the resulting GPS factors, and, in a separate multiple regression model, on GPS-Sym. In these analyses, different COVID-19 related stressors were introduced as independent variables, including change in job activity due to COVID-19, personal lockdown or quarantine status, having a loved one infected, hospitalized or deceased by COVID-19. Furthermore, the following covariates were introduced: age, gender, education, region, relational status and occupation.

Finally, we preformed descriptive statistics of the resulting GPS factors as well as overall sum score in our sample.

All analyses were performed using Stata 16® (StataCorp).

## 3. Results

### 3.1 Demographic and mental health characteristics of the sample

Demographic characteristics of the sample are reported in Table 1. A total of 18147 participants completed the questionnaire, 14448 (79.6%) women and 3699 (20.38%) men. Regional distribution was very similar to national demographical data, with 7991 (44%) of participants from Northern Italy, 4695 (25.9%) from Central Italy and 5175 (28.52%) from Southern Italy. Missing data affected 11% of observation and were thus treated by simple listwise deletion.

**Table 1.** Sample characteristics.

|  | **No. / Median (% / IQR)** |
| --- | --- |
| **Age** | 38 (23) |
| **Gender** |  |
| *Women* | 14448 (79.61) |
| *Men* | 3699 (20.38) |
| **Education** |  |
| *≤Undergraduate* | 8682 (47.84) |
| *≥Postgraduate* | 7795 (42.95) |
| *Lower education* | 1670 (9.20) |
| **Occupation** |  |
| *Housewife* | 1155 (6.36) |
| *Unemployed* | 2127 (11.72) |
| *Employed* | 10984 (60.53) |
| *Retired* | 293 (1.61) |
| *Student* | 3588 (19.77) |
| **Region** |  |
| *North* | 7991 (44.03) |
| *Centre* | 4695 (25.87) |
| *South* | 5175 (28.52) |
| *Missing* | 286 (1.58) |
| **Lockdown/quarantine** |  |
| *Lockdown* | 17486 (96.36) |
| *Quarantine* | 148 (0.8) |
| *Missing* | 513 (2.83) |
| **Working activity change** |  |
| *As usual* | 2349 (12.94) |
| *Smart-working* | 6804 (37.49) |
| *Discontinued* | 7601 (41.89) |
| *More than usual* | 672 (3.7) |
| *Missing* | 721 (3.97) |
| **Loved one's status** |  |
| *Infected* | 801 (4.41) |
| *Deceased* | 254 (1.4) |
| *Hospitalized* | 428 (2.36) |
| *Missing* | 88 (0.48) |
| **GPS-Risk/Protective Items** |  |
| *Other stressful life events* | 9325 (51.39) |
| *Poor Social Support* | 6949 (38.29) |
| *Early adverse life events* | 8637 (47.59) |
| *History of mental illness* | 5114 (28.18) |
| *Poor Resilience* | 3761 (20.73) |

Figure 1 shows the endorsement of the individual GPS symptoms. In the total sample, mean (standard deviation, SD) and median scores (interquartile range, IQR) for GPS-Sym, GPS-RP, GPS-total, PHQ-9, GAD-7 and ISI were respectively 7.2 (SD = 3.83) / 7 (IQR = 6), 1.86 (SD = 1.23) / 2 (IQR = 2), 10.6 (SD = 6.31) / 10 (IQR = 9), 8.9 (SD = 5.9) / 8 (IQR = 10) and 10.22 (SD = 7.18) / 10 (IQR = 12) (see supplementary Table S1 for further details).

**Figure 1.**
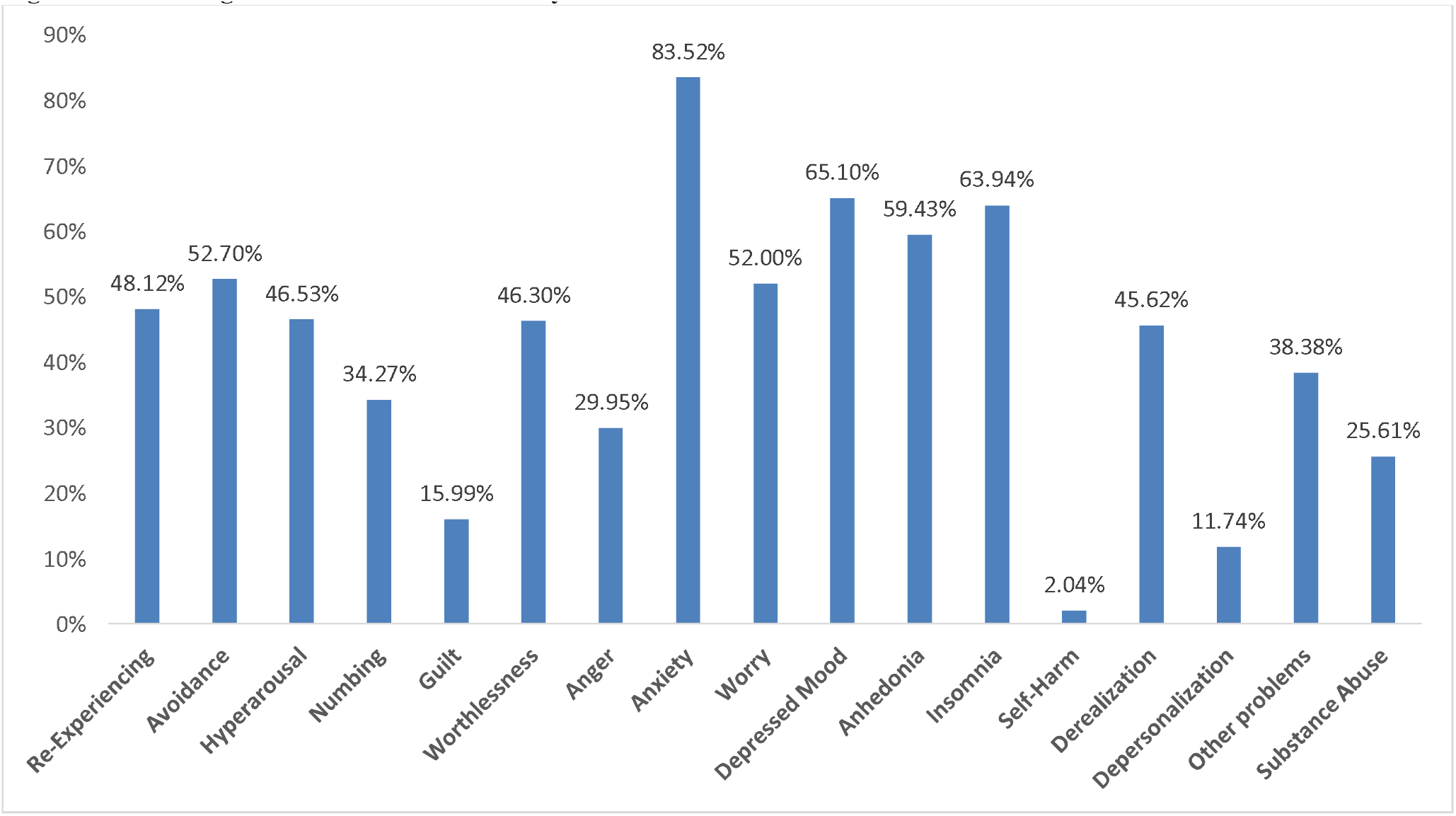
Percentage of endorsement of GPS-Sym items.

### 3.2 EFA

The Kaiser–Meyer–Olkin (KMO) measure confirmed the sampling adequacy for the analysis (KMO = 0.90) and Bartlett’s test of sphericity [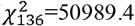 p < .001] suggested that correlations between items were suitably large, confirming the appropriateness of the analysis. Tetrachoric factor analysis revealed two contrasting solutions (Table 2).

**Table 2.**
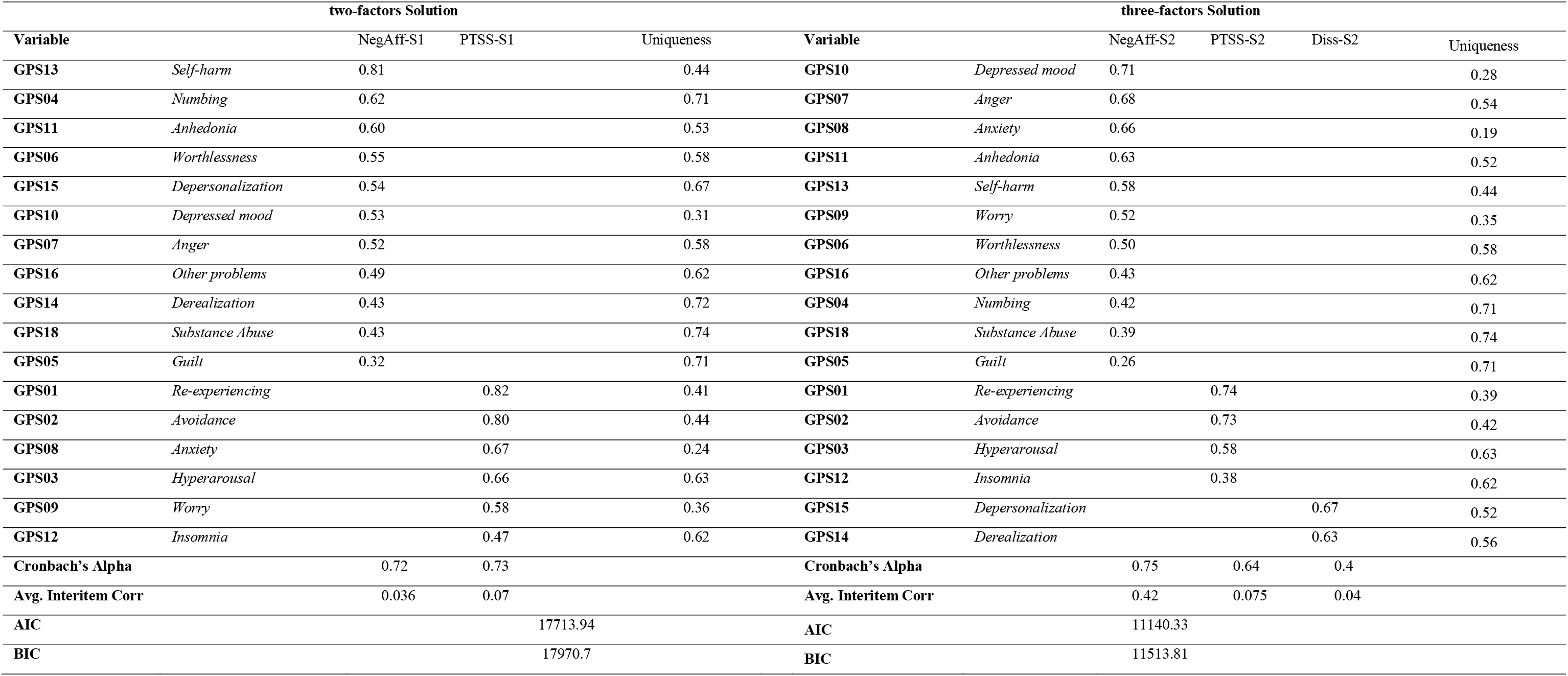
Exploratory factor analysis.

The first one (Solution 1, S1) is a two-factor solution with Re-experience, Avoidance, Anxiety, Hyperarousal, Worry and Insomnia on one factor that was termed “PTSS-S1” and a second factor termed “Negative Affect-S1” that included Self-harm, Numbing, Anhedonia, Worthlessness, Detachment, Depressed mood, Anger, Other problems, Derealization, Substance Abuse, Guilt.

The second solution (Solution 2, S2) provided three factors: the main difference was a third factor termed “Dissociation-S2” encompassing both depersonalization and derealization, while the two remaining factors (termed respectively “Negative Affect-S2” and “PTSS-S2”) were very similar S1 factors, with the exception of “Worry” loading on “Depression-S2” rather than on “PTSS-S1” factor. Furthermore, the highest loading item on Depression-S2 was depressed mood, while for Depression-S1 it was “self-Harm”.

Inspecting Eigenvalues, the two-factor solution explained 45.2% of the total variance and seemed the best solution. However, AIC and BIC criteria supported a three-factor solution that explained 48.1% of the variance. Correlations between PHQ, GAD, ISI and PSS and GPS-Sym and the factors obtained from the EFA are showed in Table 3. All correlations were significant (p< 0.001).

**Table 3.** Pairwise correlations.

|  | two-factor model |  | three-factor model |  |  | *GPS-sym* |
| --- | --- | --- | --- | --- | --- | --- |
|  | *Negative Affect* | *PTSS* | *Negative Affect* | *PTSS* | *Dissociation* |  |
| **PHQ-9** | 0.75 | 0.57 | 0.75 | 0.49 | 0.39 | 0.76 |
| **GAD-7** | 0.71 | 0.66 | 0.75 | 0.56 | 0.38 | 0.78 |
| **ISI** | 0.49 | 0.55 | 0.51 | 0.53 | 0.27 | 0.59 |
| **PSS** | 0.68 | 0.62 | 0.74 | 0.50 | 0.33 | 0.74 |

All correlation coefficients were statistically significant (p< 0.001). Results from pairwise correlations showed a strong correlation of both Depression-S1 and Depression-S2 with the PHQ (r = 0.75 in both cases) and GAD total score (r = 0.71 and r = 0.75), PTSS-S1 showed a slightly higher correlation with GAD compared with PTSS-S2 (r = 0.66 *vs*. r = 0.56). Dissociation showed moderate to small correlation with PHQ, GAD, ISI and PSS (r< 0.4).

### 3.3 CFA

Results from CFA are reported in Table 4. A one-factor solution was used as a baseline comparison, that showed acceptable fit indices (Root mean square of error approximation, RMSEA 0.058 [0.057, 0.059], Aikake’s Information criteria, AIC = 295958.409, Bayesian Information Criteria, BIC = 296355.230, Comparative Fit Index CFI = 0.862, Tucker-Lewis fit index TLI = 0.842, standardized root mean squared residual, SRMR = 0.044, coefficient of determination, CD = 0.838). Solution 1 had good fit indices, with RMSEA = 0.051 [0.05, 0.053], CFI = 0.891, TLI = 0.875, SRMR = 0.038 and CD = 0.897. Solution 2 had the best fit indices, with RMSEA = 0.047 [0.046, 0.048], CFI = 0.912, TLI = 0.897, SRMR = 0.037 and CD = 0.94.

**Table 4.**
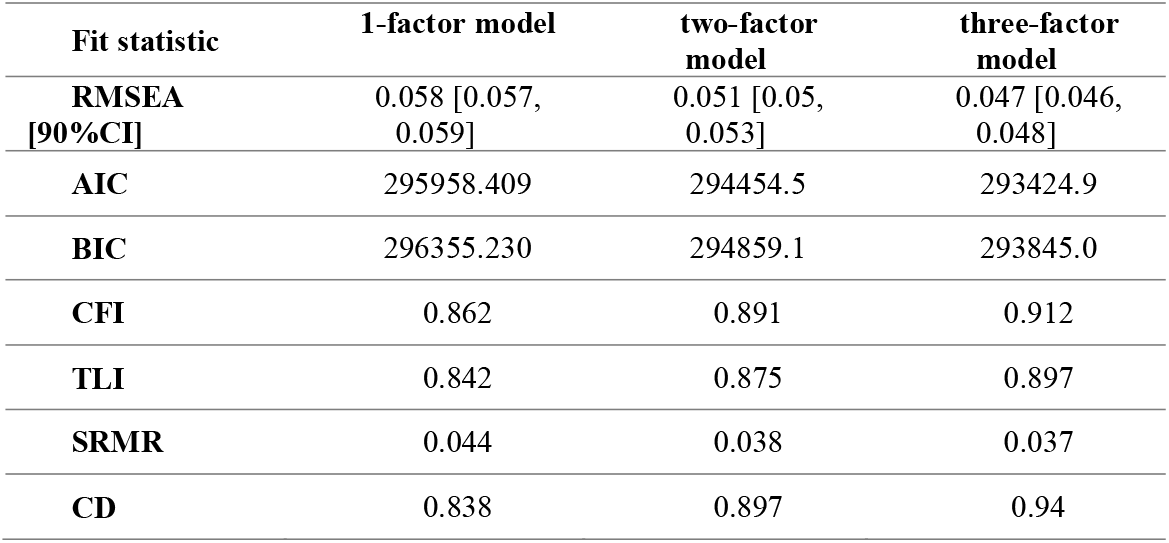
CFA Goodness-of-fit indices.

| Fit statistic | 1-factor model | two-factor model | three-factor model |
| --- | --- | --- | --- |
| **RMSEA**<br>[90%CI] | 0.058 [0.057, 0.059] | 0.051 [0.05, 0.053] | 0.047 [0.046, 0.048] |
| **AIC** | 295958.409 | 294454.5 | 293424.9 |
| **BIC** | 296355.230 | 294859.1 | 293845.0 |
| **CFI** | 0.862 | 0.891 | 0.912 |
| **TLI** | 0.842 | 0.875 | 0.897 |
| **SRMR** | 0.044 | 0.038 | 0.037 |
| **CD** | 0.838 | 0.897 | 0.94 |

### 3.4 Trauma spectrum in the population and its relationship with general and COVID-related risk factors

The raw and averaged scores of the three resulting GPS factors were calculated and reported in Table 5. Scores were averaged in order to be more easily comparable between each other. In our sample, participants endorsed on the average 4.52/11 Negative affect symptoms, 2.11/4 PTSS and 0.57/2 Dissociative symptoms.

**Table 5.**
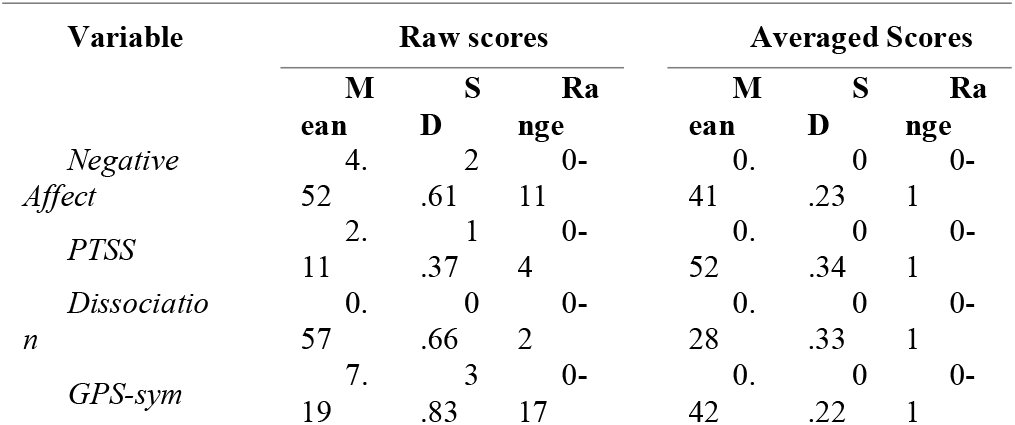
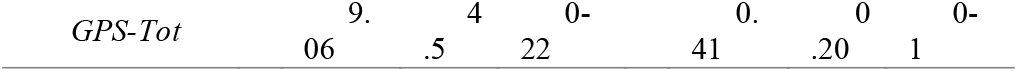

The relationship between GPS factors Negative Affect, PTSS and Dissociation and a number of risk factors was assessed by seemingly unrelated linear regression (sureg) analysis (see Table 6). Parallel to the sureg model, a multiple regression model was fitted with the same independent variables on GPS-Sym. Due to missingness, final sureg model was estimated on data from 17231 respondents. In order to have more easily interpretable results, dependent variables scores were standardized. Younger age was more strongly associated with Negative Affect compared to PTSS or Dissociation (b = 0.23 [0.21, 0.24] compared to b = 0.05 [0.03, 0.07] and b = 0.05 [0.04, 0.07]. Female gender showed a larger association with PTSS (b = 0.42 [0.38, 0.45]) compared to Negative Affect (b = 0.24 [0.21, 0.27]) or Dissociation (b = 0.20 [0.16, 0.24]). Compared to being in lockdown, being under quarantine because infected or exposed to someone infected was associated with both Negative Affect and PTSS (b = 0.25 [0.10, 0.39] and b = 0.22 [0.06, 0.38]). Participants from Southern Italy had selectively higher PTSS symptoms than participants from Northern or Central Italy (b = 0.18 [0.15, 0.22]). Being a housewife was associated with Negative Affect and PTSS (b = 0.11 [0.05, 0.16] and b = 0.15 [0.08, 0.21]. Being a student was associate with lower PTSS (b = –0.12 [-0.17, –0.08]). Having experienced any stressful life event due to COVID-19 was associated Negative Affect, PTSS and Dissociation (b = 0.22 [0.19, 0.24], b = 0.15 [0.12, 0.18] and b = 0.12 [0.09, 0.15]). Working more than usual was associated with PTSS and dissociation (b = 0.19 [0.10, 0.28] and b = 0.10 [0.01, 0.19]). Having a loved one deceased by COVID19 was associated with Negative Affect and PTSS (b = 0.15 [0.05, 0.26] and b = 0.25 [0.13, 0.37]) Having a loved one hospitalized was associated with PTSS (b = 0.14 [0.05, 0.24]). GPS-RP was associated with Negative Affect, PTSS and Dissociation (b = 0.30 [0.29, 0.31], b = 0.16 [0.15, 0.18] and b = 0.15 [0.13, 0.16]).

**Table 6.**
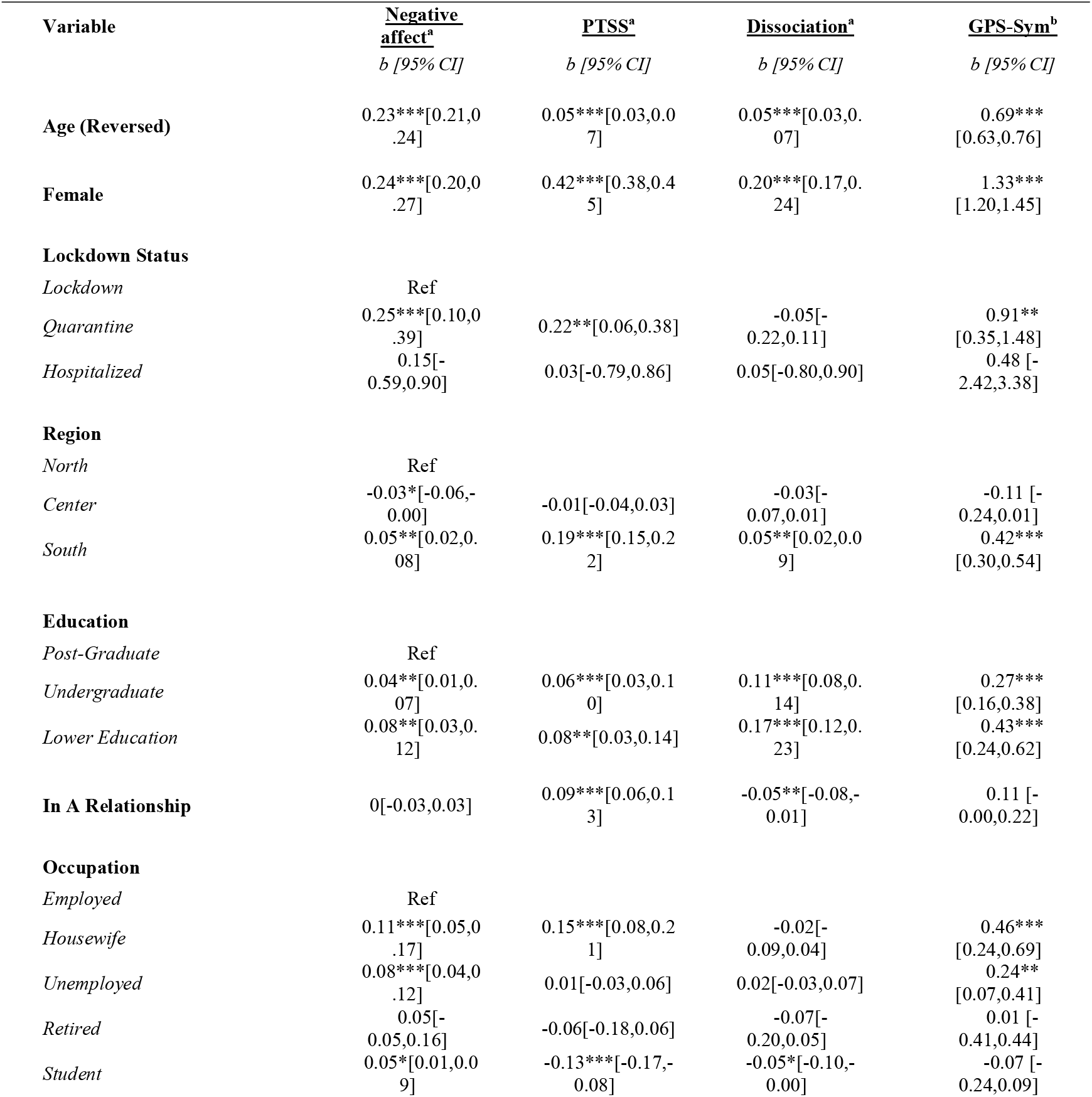
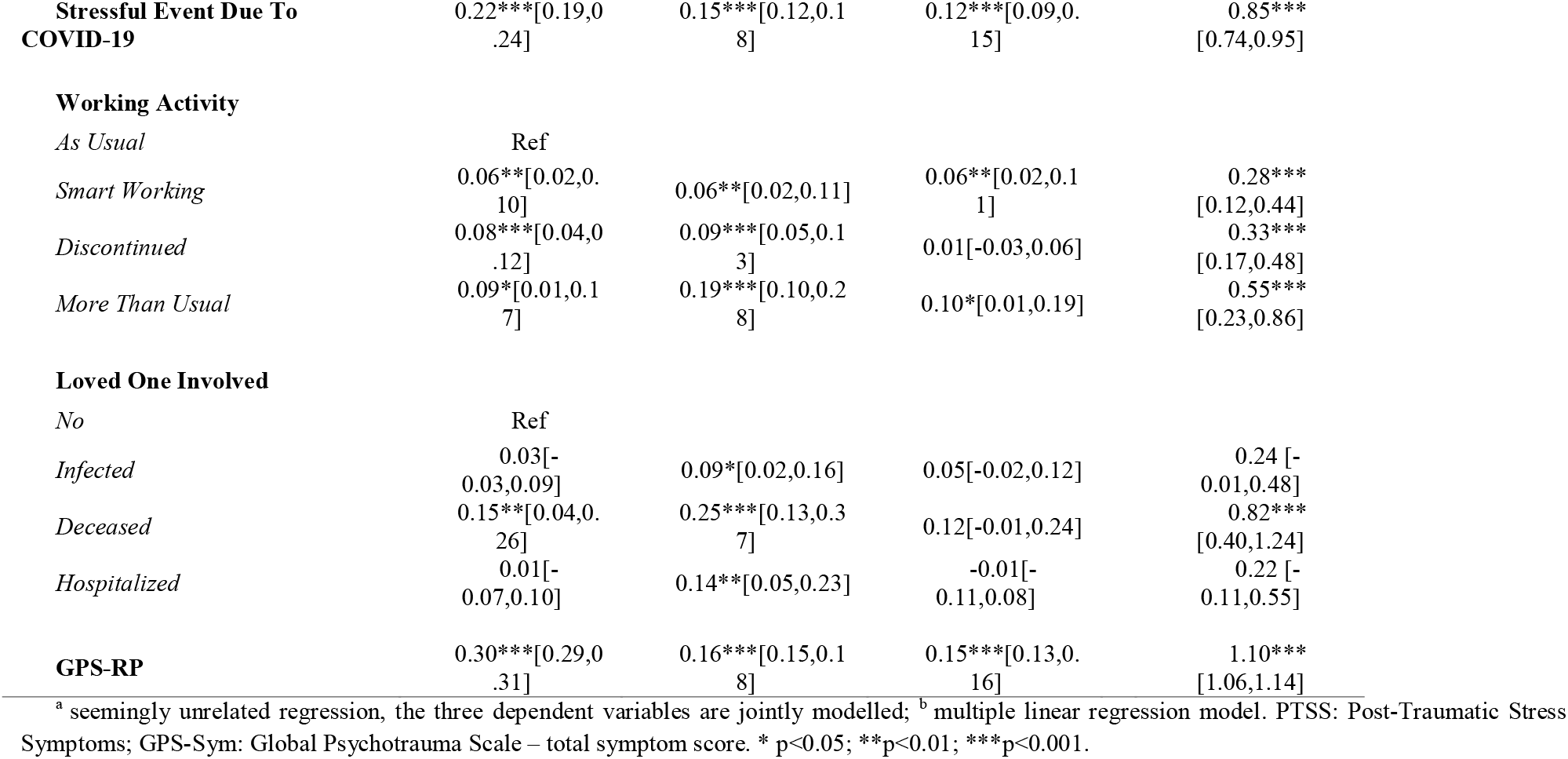
Demographic and risk/protective factors as predictors of GPS scores. PLEASE INDICATE WHAT ’s REPRESENT

## 4. Discussion

This study had the objective to explore the factor structure of the Italian version of the Global Psychotrauma Screen (GPS), which assesses the wide range of traumatic spectrum symptoms, in a large web-based study carried out during the COVID-19 pandemic. Secondly, we explored the association between traumatic spectrum symptoms and GPS and COVID-19 related risk factors. This study is an addition to the field of stress and trauma related disorders, as it allows to detail how a wide range of traumatic symptoms may be associated with the exposure to a pandemic and lockdown measures in the general population. To the best of our knowledge, this is the first report on post-traumatic symptoms in the Italian population since the COVID-19 outbreak.

The GPS is brief screening tool but at the same time taps a wide range of post-traumatic symptoms. Indeed, a broad spectrum of symptoms was endorsed, with only the Self-harm item being endorsed by a small minority, an item that may be expected more after severe and or long-lasting interpersonal violence. GPS-Total Score was within the same range of that found in a larger study on child maltreatment (Olff et al., 2020). Although a one-factor model showed acceptable fit, our psychometric analyses support that a three-factor structure of post-traumatic symptoms has superior fit. In this model, one factor encompasses depressive and anxious symptoms as well as anger and irritability, and was termed “Negative Affect”; a second factor includes core post-traumatic symptoms (“PTSS”), such as re-experiencing, avoidance and hyperarousal, and sleep disturbances; a third factor includes “Dissociative symptoms”, i.e. depersonalization and derealization.

The factor structure emerged from these study needs to be discussed in the light of recent taxonomic advances in stress-related disorders proposed in the International Classification of Diseases – 11^th^ edition (ICD-11) and the diagnostic and Statistical Manual of Mental Disorders 5^th^ edition (DSM 5).

The first factor, i.e. “Negative Affect”, includes depressed mood, irritability, anxiety as well as suicidal ideation, sense of worthlessness, guilt, and Substance Abuse. This factor partially overlaps with DSM 5 criterion D “Negative alterations in mood and cognition”. Furthermore, the “Negative Affect” factor partially overlaps with the “disturbance in self organization” (DSO) that characterizes complex-PTSD in ICD-11 (Cloitre et al., 2013; Karatzias et al., 2017; Shevlin et al., 2018).

The factor “PTSS” includes the classic PTSD core symptoms, i.e. hyperarousal, re-experiencing and avoidance similar to those in the ICD-11 PTSD cluster, plus sleep problems.

Finally, in the GPS, dissociative symptoms load on a separate factor, coherently with the DSM 5 dissociative specification (Hansen et al., 2017b; Longo et al., 2019; Rossi et al., 2019). The fact that dissociation loads onto a separate factor supports the Subtype model of the relationship between PTSD and dissociation (Hansen et al., 2017b; Ross et al., 2018).

The inclusion of cognitive and affective symptoms in the PTSD criteria by DSM 5 has produced a large number of different hypothesized latent factor structures of PTSD, with the result of an increased confusion in stress-related disorders taxonomy (Hansen et al., 2015). At the same time, ICD-11 diagnostic criteria are associated with increased diagnostic accuracy, reducing comorbidities with other traumatic responses (Cloitre et al., 2013; Hansen et al., 2015, 2017a). Furthermore, ICD-11 criteria are associated with a much more limited number of proposed latent structures in the literature (Hansen et al., 2015). Our data suggest that core PTSD symptoms are separated from affective and cognitive symptoms, roughly resembling the ICD-11 organization of PTSD and cPTSD symptoms. Our data add to the theory that affective symptoms are something separated from core PTSS and could contribute to a more severe PTSD clinical picture, or a distinct disorder such as cPTSD. Another relevant aspect of negative affective symptoms is its high centrality in DSM-5 based networks of PTSS, that is thought to underlie some mechanisms underpinning high comorbidities with other mental disorders (Birkeland et al., 2020).

Following factor analyses, we assessed how trauma-related symptoms were associated with several risk factors and covariates, including demographic variables, a history of early trauma or mental illness or missing a supportive environment and COVID-19 specific factors as well as a risk-protection index included in the GPS, such as having.

With regard to demographic variables, women were more likely to endorse the GPS total as well as the three symptoms-clusters, particularly the PTSS factor, while young age was particularly associate with Negative Affect. This is in line with previous PTSD research (De Vries and Olff, 2009; Dückers and Olff, 2017; Olff, 2017) and with other studies from China that highlight a higher vulnerability for stress-related symptoms in women (Liu et al., 2020; Qiu et al., 2020; Wang et al., 2020). Other demographic variables associated with a higher GPS scores were lower education level and being unemployed or a housewife, as also found in China (Wang et al., 2020). GPS risk and protective factors (including poor social support and a history of trauma and psychiatric disorders or low sense of resilience) were associated with higher symptom profiles.

COVID-19 related stressful events were also associated with the GPS total symptoms and the three factors. Being under quarantine because of being infected or exposed to contagion and having lost a loved one due to COVID-19 were associated with higher levels of PTSS and Negative Affect. COVID-19 related stressful events were correlated with the three symptoms clusters. These findings confirm a relevant impact of COVID-19 related events on mental health, even after adjusting for pre-existing risk factors included in the GPS-RP such as early traumatic experiences, previous mental illness or poor social support. These results warrant a close monitoring of the evolution of stress-related symptoms in the general population over time and support the urge to enforce mental health interventions.

This study has several limitations. It is not a representative population sample. Social-network sampling strategy has its pros and cons. In the context of the COVID-19 pandemic, it was essential to collect a large sample in a very short time, as a part of a long-term monitoring programme of mental health outcomes in the general population (Rossi et al., 2020b). However, this web-based survey may have introduced a number of potential biases, including self-selection bias as suggested by the large disproportion in the gender ratio and the unusually high rate of self-reported lifetime prevalence of a history of mental illness or psychiatric/psychological treatment at around 28%. It might indicate that an already more vulnerable group in the population is more inclined to fill in a survey on mental health effects of the pandemic, and thus lead to an overestimation the prevalence of traumatic-spectrum symptoms. Furthermore, only a limited number of instruments could be included in the survey. Ideally a larger battery would have been included to assess concurrent and divergent validity of the GPS. Also, self-report instruments inherently introduce measuring biases, especially in the absence of normative data. Another major limitation is the absence of clinical interviews or other normative cut-offs, that would have allowed to estimate a prevalence of PTSD or clinically relevant PTSS. A strength of the study is its large sample size, and the timely data collection around the peak of the COVID-19 outbreak providing a unique set of data on the Italian effects of the pandemic in its early phases.

## 5. Conclusion

This is the first study to address in detail the traumatic spectrum related to COVID-19 in Italy, and to provide a first validation of the Italian GPS, showing the psychometric properties of a novel screening tool for trauma-related symptoms. The GPS captured the wide range of symptoms with a three-factor model best explaining the symptoms: 1. Negative Affect; 2. Core post-traumatic stress symptoms (PTSS); 3. Dissociative symptoms. Our findings suggest that the COVID-19 pandemic and related lockdown measures in Italy are seriously affecting general population’s mental health with a wide range of trauma spectrum symptoms being endorsed.

## Data Availability

data available upon request from the authors

## Acknowledgements

This study was supported by Territori Aperti, a project funded by the Fondo Territori Lavoro e Conoscenza of the Confederazione Generale Italiana del Lavoro, the Confederazione Italiana Sindacati Lavoratori, and the Unione Italiana del Lavoro.

---

**Table S1:** GPS scores by demographics.

|  | Negative Affect | PTSS | Dissociation | GPS-Sym | GPS-Tot |
| --- | --- | --- | --- | --- | --- |
|  | Mean (SD) | Mean (SD) | Mean (SD) | Mean (SD) | Mean (SD) |
| **Female** | 0.42 (0.23) | 0.56 (0.33) | 0.3 (0.33) | 7.56 (3.74) | 0.43 (0.20) |
| **Male** | 0.34 (0.24) | 0.38 (0.32) | 0.22 (0.3) | 5.76 (3.82) | 0.33 (0.20) |
| **18-35** | 0.47 (0.23) | 0.53 (0.33) | 0.30 (0.34) | 8.01 (3.72) | 0.45 (0.20) |
| **36-50** | 0.38 (0.22) | 0.53 (0.34) | 0.27 (0.32) | 6.91 (3.75) | 0.40 (0.20) |
| **51-65** | 0.31 (0.22) | 0.48 (0.34) | 0.25 (0.31) | 5.90 (3.78) | 0.34 (0.20) |
| **>65** | 0.23 (0.20) | 0.44 (0.32) | 0.14 (0.27) | 4.66 (3.44) | 0.28 (0.19) |
| **North** | 0.40 (0.24) | 0.51 (0.34) | 0.28 (0.32) | 7.13 (3.86) | 0.40 (0.20) |
| **Centre** | 0.39 (0.23) | 0.50 (0.33) | 0.27 (0.32) | 6.91 (3.74) | 0.39 (0.20) |
| **South** | 0.42 (0.23) | 0.57 (0.33) | 0.30 (0.34) | 7.54 (3.82) | 0.42 (0.20) |
| **Post-Graduate** | 0.39 (0.22) | 0.50(0.34) | 0.25 (0.31) | 6.87 (3.72) | 0.39 (0.19) |
| **Under-graduate** | 0.42 (0.24) | 0.53 (0.34) | 0.30 (0.33) | 7.40 (3.86) | 0.42 (0.20) |
| **Lower education** | 0.43 (0.25) | 0.55 (0.34) | 0.32 (0.34) | 7.61 (4.00) | 0.44 (0.21) |
| **Housewife** | 0.40 (0.22) | 0.61 (0.34) | 0.29 (0.33) | 7.54 (3.72) | 0.43 (0.20) |
| **Unemployed** | 0.46 (0.23) | 0.57 (0.33) | 0.32 (0.34) | 8.10 (3.77) | 0.47 (0.20) |
| **Employed** | 0.37 (0.23) | 0.51 (0.34) | 0.27 (0.32) | 6.74 (3.78) | 0.38 (0.20) |
| **Retired** | 0.27 (0.24) | 0.42 (0.33) | 0.19 (0.30) | 5.11 (4.06) | 0.30 (0.21) |
| **Student** | 0.49 (0.22) | 0.51 (0.33) | 0.30 (0.34) | 8.11 (3.71) | 0.45 (0.19) |

